# Prevalence of sarcopenia and frailty in mobility-limited older adults: the cross-sectional BIOFRAIL cohort study

**DOI:** 10.64898/2026.11.18.26368392

**Authors:** Pernille Hansen, Hanne Nygaard, Rikke Stefan Kamper, Martin Schultz, Jesper Ryg, Per Aagaard, Flemming Dela, Charlotte Suetta

## Abstract

**Background:** Sarcopenia and frailty pose considerable risks to older adults due to their close association with reduced physical performance, increased risks of falls, and mortality. Despite this, knowledge about the prevalence in mobility-limited older adults remains scarce. Therefore, we aimed to identify the prevalence and co-prevalence of sarcopenia and frailty in mobility-limited older adults.

**Methods:** This cross-sectional study included patients aged ≥65 years referred to a geriatric outpatient clinic for fall assessment. Sarcopenia risk was screened using SARC-F and defined according to the European Working Group on Sarcopenia in Older People criteria; frailty was evaluated using the Clinical Frailty Scale. Muscle strength was assessed by handgrip strength and the 30-second sit-to-stand test. Skeletal muscle index was determined using bioelectrical impedance analysis (Inbody770). Gait speed was assessed over 6 meters.

**Results:** A total of 375 patients (mean age 79.7 ± 6.1 years; 65% female) were included, with 28.0% demonstrating low HGS and 36.8% showing low STS performance, while only 13.9% demonstrated concurrent impairments in both tests. Low SMI was observed in 25.1% of the patients. The prevalence of sarcopenia was 14.1%, frailty 23.7%, and their co-prevalence 7.2%.

**Conclusions:** In mobility-limited older adults referred for fall assessment, sarcopenia and frailty were common conditions showing only limited overlap. The low co-prevalence between HGS and STS underscores the importance of using multiple strength assessments, while the substantial prevalence of low muscle mass highlights the need for routine muscle mass evaluation regardless of muscle strength status.

**Trial registration:** NCT05795556

## 1. Background

In recent decades, the proportion of older people and their life expectancy has risen, a trend that is projected to persist (Denmark Statistics, 2021). In parallel with the increasing aging population, the number of frail older people with age-related diseases and disabilities also increases (Clegg et al., 2013). Frailty and sarcopenia are two critical conditions affecting older adults, often leading to a decline in physical health and quality of life. While many studies focus on either frailty or sarcopenia independently (Bauer & Sieber, 2008), only few studies have addressed both conditions despite the substantial overlap and the compounded risks they pose (Davies et al., 2018; Nygaard et al., 2024).

Sarcopenia is characterized by progressive and differential loss of muscle strength and muscle mass (Cruz-Jentoft et al., 2010) and patients with sarcopenia have a higher risk of disability, falls and fall-related injuries (Bischoff-Ferrari et al., 2015a; Schaap et al., 2018), reduced independency (Malmstrom et al., 2016), and low quality of life (Beaudart et al., 2017). In 2010, the European Working Group on Sarcopenia in Older People (EWGSOP) proposed a practical clinical definition including consensus diagnosis criteria for sarcopenia (Cruz-Jentoft et al., 2010) with an updated version in 2018 (EWGSOP2) (Cruz-Jentoft et al., 2019).

Frailty is characterized by a decline in multiple physiological systems, resulting in increased vulnerability to various stressors (Chen et al., 2014; Clegg et al., 2013; Howlett & Rockwood, 2013; Morley, 2014). The condition is often displayed by reduced physical ability, high comorbidity burden, dependency of others to manage activities of daily living (ADL), and reduced life expectancy (Clegg et al., 2013; Dent et al., 2016). Even though frailty is a well-recognized syndrome (Chen et al., 2014), particularly in geriatric populations, no universally accepted definition of the syndrome exists (Morley, 2014). Instead, various definitions and criteria have been proposed (Dent et al., 2016; Oviedo-Briones et al., 2022; Pérez-Zepeda et al., 2016). The Clinical Frailty Scale (CFS) is a clinical judgement-based tool developed for the Canadian Study of Health and Aging (Rockwood et al., 2005) that is based on scores from different domains such as comorbidity, physical performance, ADL dependency, and cognition (Rockwood et al., 2005). The CFS has been translated into multiple languages including Danish (Nissen et al., 2020) and evaluated in numerous clinical studies and reviews concluding that the CFS is a valid, simple, and feasible tool for frailty screening (Belga et al., 2016; Church et al., 2020).

In the scientific literature, a substantial overlap regarding characteristics, etiological factors, and risk of adverse outcomes between frailty and sarcopenia is described (Bischoff-Ferrari et al., 2015b; Chen et al., 2014; Clegg et al., 2013; de Buyser et al., 2016; Jeejeebhoy, 2012). This overlap suggests that the two geriatric conditions share common underlying mechanisms, such as chronic inflammation, hormonal changes, and nutritional deficiencies (Jeejeebhoy, 2012), altogether resulting in reduced mechanical muscle function (Aagaard et al., 2010).

Importantly, reductions in muscle strength, muscle mass, and gait speed are not only used as diagnostic markers for sarcopenia but are also recognized as key risk factors for falls and indicators of frailty in older people (Cruz-Jentoft et al., 2019; Tieland et al., 2018). However, despite the clear association between sarcopenia, frailty, and falls (Yeung et al., 2019), there is a notable gap in the literature regarding the prevalence of sarcopenia and frailty, specifically among fallers and mobility-limited older adults. Only few studies have assessed the prevalence of sarcopenia and frailty in older people with a previous history of falls (Barnes et al., 2018; Christensen et al., 2018; Church et al., 2020; Reijnierse et al., 2016) but to the best of our knowledge, no previous studies so far have investigated the prevalence of sarcopenia in populations at high risk of falling based on the updated EWGSOP2 criteria. Improving our understanding of the prevalence of sarcopenia and frailty, as well as the overlap between these two conditions among a high-risk population is important for implementing targeted interventions to prevent falls and mitigate their consequences in old age.

The purpose of the present study, therefore, was to identify the prevalence and co-prevalence of sarcopenia and frailty in mobility-limited older adults.

## 2. Material & Methods

### 2.1. Study design and Setting

The BIOFRAIL Study is a cross-sectional cohort study of patients referred to the geriatric outpatient clinic at Copenhagen University Hospital, Herlev and Gentofte, Denmark, for fall assessment from September 2021 to June 2023. All patients referred to the clinic aged 65 years and above were eligible for the study. Exclusion criteria were (i) age < 65 years, (ii) severe dementia, (iii) severe communicative problems, (iv) inability to provide informed consent for study participation or inability to follow the test protocol, and (v) inability to walk independently without personal support (walking aids allowed).

All enrolled patients provided informed written consent. The study was approved by the local Ethics Committee of Copenhagen and Frederiksberg (H-20057620), the Danish Data Protection Agency (P-2021-619), and registered at Clinicaltrials.gov (14. February 2022, NCT05795556). All experimental procedures were performed in accordance with the Declaration of Helsinki. This study was conducted and reported in accordance with the STROBE (Strengthening the Reporting of Observational Studies in Epidemiology) guidelines (von Elm et al., 2014) to ensure transparency and completeness in the reporting of study design, data collection, and analysis.

### 2.2. Physical measurements

Height (m) was measured without shoes, to the nearest 0.1 cm and weight (kg) was assessed with light clothes on, to the nearest 0.1 kg. Subsequently body mass index (BMI) was calculated (kg/m²).

### 2.3. Frailty

CFS was used to assess the degree of frailty on a scale from 1 (very fit) to 9 (terminally ill), based on clinical judgement and consideration of a patient’s functional status, comorbidities, and the degree of dependency in ADL-activities. Scores ≥5 specify the presence of frailty (Rockwood et al., 2005).

### 2.4. Sarcopenia

EWGSOP2 guidelines were used to identify sarcopenia (Cruz-Jentoft et al., 2019). According to these guidelines, sarcopenia was considered evident when both low muscle strength and low muscle mass were present. Low muscle strength was present if the patients exhibited low values in either or both handgrip strength (HGS) or sit-to-stand (STS) performance. Severe sarcopenia was evident when both low muscle strength, low muscle mass, and low habitual gait speed were present (Cruz-Jentoft et al., 2019).

The SARC-F questionnaire (Malmstrom et al., 2016) was used to screen patients for self-reported signs of sarcopenia, which include self-perceived habitual strength, physical performance, and number of falls within the last year. Fall incidents were classified as either (i) no falls within the last 12 months, (ii) 1-3 falls, or (iii) 4 or more falls according to SARC-F (Malmstrom et al., 2016). As proposed by Malmstrom (Malmstrom et al., 2016), the risk of sarcopenia was defined as SARC-F ≥4.

### 2.5. Muscle strength

HGS was assessed using a hand-held dynamometer (Jamar Smart, Sammons Preston Rolyan, Chicago, Illinois, USA), where a minimum of three attempts were performed for the dominant hand, and the highest value was selected for statistical analysis. The cut-off values for low HGS were <27 kg and <16 kg, for male and female, respectively (Cruz-Jentoft et al., 2019). Sit-to-stand performance was assessed using the 30-sec STS chair test (Jones et al., 1999), with a low STS performance defined as ≤8 repetitions (Bruun et al., 2019; Rikli & Jones, 2013).

### 2.6. Muscle mass

Muscle mass was assessed using direct segmental 8-electrode multi-frequency bioelectrical impedance analyses (BIA) (Inbody770 and InbodyS10; Biospace Co., Seoul, Korea) and reported as skeletal muscle index (SMI) (Appendicular skeletal muscle mass (sum of the muscle mass of the arms and legs) adjusted for the height^2^).

As muscle mass measured with BIA appears to slightly overestimate similar measures obtained by DXA, data were adjusted using the equation derived by Kristensen and colleagues in older people (Kristensen et al., 2025):

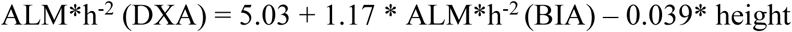

Critical cut-off values for low muscle mass were <7.0 kg/m^2^ and <5.5 kg/m^2^, for male and female, respectively (Cruz-Jentoft et al., 2019).

### 2.7. Physical performance

Horizontal gait speed (GS) was assessed as habitual (self-chosen) and maximal GS, respectively, recorded during horizontal walking over a 6-meter indoor course (Bohannon, 1997). Habitual walking aids were allowed. GS was computed as the 6-meter distance divided by the elapsed time of the two attempts in each condition (Bohannon, 1997; Sustakoski et al., 2016). Critical cut-off values for low GS were defined as ≤0.8 m/s (Cruz-Jentoft et al., 2019).

### 2.8. Assessments related to patient characteristics

The Short Nutritional Assessment Questionnaire (SNAQ) was used to determine malnutrition risk (Kruizenga et al., 2005). Patients scoring below 2 points were observed as well-nourished, a score of 2 indicated moderate malnutrition, and scores of 3 or higher indicated severe malnutrition. Depression severity was measured using the 15-item Geriatric Depression Scale (GDS-15). A score of 0-4 indicated no depression, scores of 5-8 suggested mild depression, 9-11 pointed to moderate depression, and scores of 12 or above represented severe depression (Djernes et al., 2004; Shin et al., 2019).

### 2.9. Statistical analysis

All statistical analysis was performed in IBM SPSS Statistics. Descriptive statistics were conducted to determine mean values, variance, and prevalence rates of given outcome variables, which were presented as mean ± standard deviation (SD), median with interquartile range [IQR], or relative frequency (%) unless otherwise stated.

All continuous data were visually inspected in residual plots and histograms and were assessed for normality. Independent *t*-tests, Mann-Whitney U test, and chi-squared test were used to investigate potential differences between sexes. Statistical significance was set at p≤0.05 (two-tailed).

## 3. Results

A total of 375 patients with complete records of HGS, STS, SMI, and CFS were included in the study, mean ± SD age 79.7 ± 6.1 and 65% female. The characteristics of the study population are listed in Table 1.

**Table 1.**
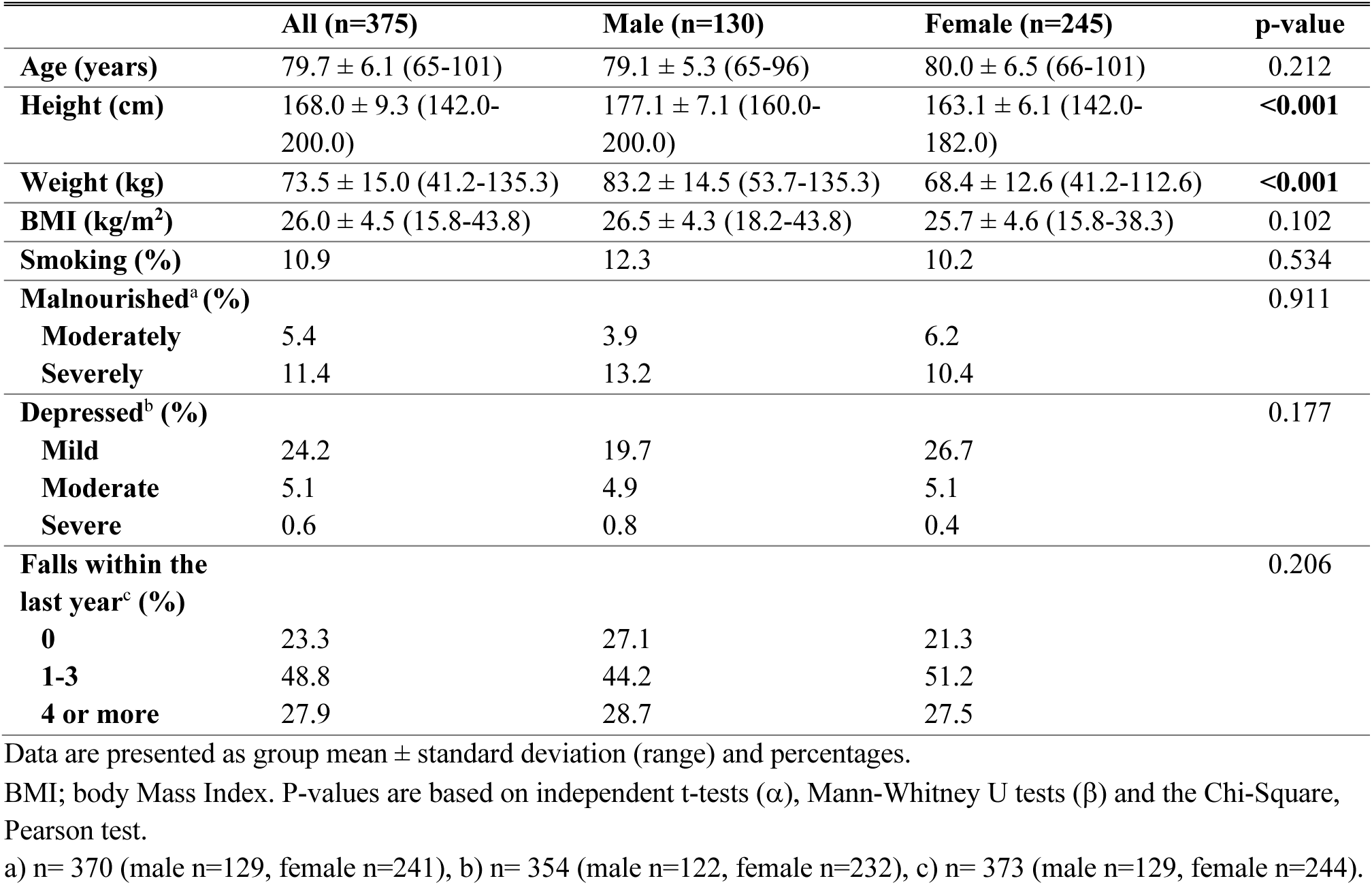
Characteristics of the study population.

|  | **All (n=375)** | **Male (n=130)** | **Female (n=245)** | **p-value** |
|---|---|---|---|---|
| **Age (years)** | 79.7 ± 6.1 (65-101) | 79.1 ± 5.3 (65-96) | 80.0 ± 6.5 (66-101) | 0.212 |
| **Height (cm)** | 168.0 ± 9.3 (142.0-200.0) | 177.1 ± 7.1 (160.0-200.0) | 163.1 ± 6.1 (142.0-182.0) | **<0.001** |
| **Weight (kg)** | 73.5 ± 15.0 (41.2-135.3) | 83.2 ± 14.5 (53.7-135.3) | 68.4 ± 12.6 (41.2-112.6) | **<0.001** |
| **BMI (kg/m<sup>2</sup>)** | 26.0 ± 4.5 (15.8-43.8) | 26.5 ± 4.3 (18.2-43.8) | 25.7 ± 4.6 (15.8-38.3) | 0.102 |
| **Smoking (%)** | 10.9 | 12.3 | 10.2 | 0.534 |
| **Malnourished<sup>a</sup> (%)** |  |  |  | 0.911 |
| **Moderately** | 5.4 | 3.9 | 6.2 |  |
| **Severely** | 11.4 | 13.2 | 10.4 |  |
| **Depressed<sup>b</sup> (%)** |  |  |  | 0.177 |
| **Mild** | 24.2 | 19.7 | 26.7 |  |
| **Moderate** | 5.1 | 4.9 | 5.1 |  |
| **Severe** | 0.6 | 0.8 | 0.4 |  |
| **Falls within the last year<sup>c</sup> (%)** |  |  |  | 0.206 |
| **0** | 23.3 | 27.1 | 21.3 |  |
| **1-3** | 48.8 | 44.2 | 51.2 |  |
| **4 or more** | 27.9 | 28.7 | 27.5 |  |

Data are presented as group mean ± standard deviation (range) and percentages.
BMI; body Mass Index. P-values are based on independent t-tests (α), Mann-Whitney U tests (β) and the Chi-Square, Pearson test.
a) n= 370 (male n=129, female n=241), b) n= 354 (male n=122, female n=232), c) n= 373 (male n=129, female n=244).

Within the last year, 76.7% of the patients had fallen (with no differences between sexes), of which 48.8% had experienced 1-3 falls (males: 44.2% vs females: 51.2%), whereas 27.9% had experienced 4 or more falls (males: 28.7% vs females: 27.5%). According to SARC-F, the prevalence of patients at risk of sarcopenia was 48% (males: 38.8% vs females: 52.9%, p=0.009) with a median score of 3 [2-6] (males: 3 [1-5.5] vs females: 4 [2-6], p=0.021) (Table 2).

**Table 2.** Functional characteristics for all patients.

|  | **All (n=175)** | **Male (n=130)** | **Female (n=245)** | **p-value** |
|---|---|---|---|---|
| **SARC-F**<sup>a</sup> | 3.8 ± 2.6;<br>3 [2-6] | 3.4 ± 2.7;<br>3 [1-5.5] | 4.1 ± 2.5;<br>4 [2-6] | **0.021**<sup>β</sup> |
| **Low SARC-F** | 48.0% | 38.8% | 52.9% | **0.009** |
| **Hand grip strength (HGS) (Kg)** | 23.4 ± 9.2;<br>21.6 [17.0-28.9] | 32.4 ± 8.4;<br>31.7 [27.1-38.6] | 18.6 ± 5.2;<br>18.6 [15.0-22.0] | **<0.001**<sup>β</sup> |
| **Low HGS** | 28.0% | 23.8% | 30.2% | 0.192 |
| **Sit-to-stand performance (STS) (reps)** | 9.7 ± 5.2;<br>10 [7-13] | 9.8 ± 6.1;<br>11[ 7-13.3] | 9.6 ± 4.7;<br>10 [7-13] | 0.545<sup>β</sup> |
| **Low STS** | 36.8% | 36.9% | 36.7% | 0.971 |
| **Skeletal Muscle Index (SMI) (Kg/m<sup>2</sup>)** | 6.6 ± 1.1;<br>6.6[ 5.8-7.2] | 7.6 ± 1.0;<br>7.6 [7.0-8.1] | 6.1 ± 0.8;<br>6.1 [5.5-6.7] | **<0.001**<sup>α</sup> |
| **Low SMI** | 25.1% | 27.7% | 23.7% | 0.393 |
| **6-m habitual gait speed**<sup>b</sup> **(GS) (m/s)** | 0.87 ± 0.26;<br>0.86 [0.69-1.05] | 0.89 ± 0.25;<br>0.88 [0.74-1.06] | 0.86 ± 0.26;<br>0.86 [0.69-1.06] | 0.398<sup>α</sup> |
| **Low habitual GS** | 40.3% | 33.9% | 43.8% | 0.065 |
| **6-m maximal gait speed**<sup>b</sup> **(GS) (m/s)** | 1.16 ± 0.37<br>1.15 [0.90-1.40] | 1.23 ± 0.38;<br>1.22 [0.98-1.43] | 1.13 ± 0.35;<br>1.12 [0.87-1.36] | **0.028**<sup>α</sup> |
| **Low maximal GS** | 16.3% | 13.4% | 18.0% | 0.270 |
| **Sarcopenia** | 14.1% | 15.4% | 13.5% | 0.612 |
| **Frailty** | 23.7% | 22.3% | 24.5% | 0.636 |

Data are presented as group mean ± standard deviation, median [Q1-Q3], and percentages.
SARC-F, 5-item questionnaire for sarcopenia screening. Low SARC-F defined as scores ≥4. Low HGS defined as values <27 kg and <16 kg, for men and women, respectively. Low STS performance defined as values ≤8 repetitions. Low SMI defined as values <7.0 kg/m<sup>2</sup> and <5.5 kg/m<sup>2</sup>, for men and women, respectively. Low GS defined as values ≤0.8 m/s. Sarcopenia was defined as low HGS or/and low STS and low SMI (see text for details). P-values are based on independent t-tests (α), Mann-Whitney U tests (β) and the Chi-Square, Pearson test.
a) n= 373 (male n=129, female n=244) and b) n= 362 (male n=127, female n=235).

The prevalence of sarcopenia was 14.1% (males: 15.4% vs females: 13.5%, p=0.612) and the prevalence of severe sarcopenia was 8.3% (males: 11% vs females: 6.8%, p=0.165).

The prevalence of frailty was 23.7% (males: 22.3% vs females: 24.5%, p=0.636), whereas the co-prevalence of sarcopenia and frailty was 7.2% (males: 7.7% vs females: 6.9%, p=0.788) (Figure 1).

**Figure 1.**
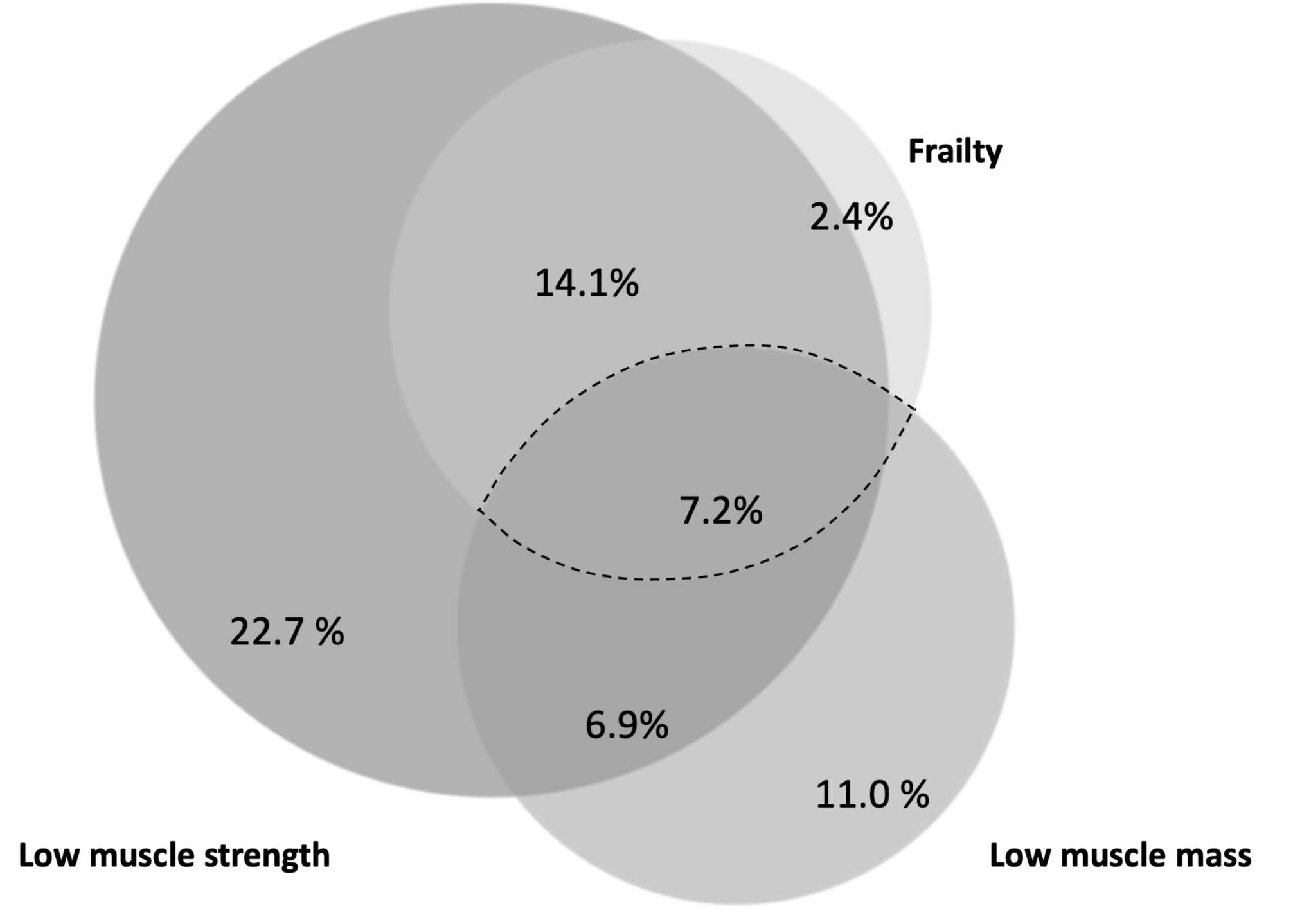
Venn diagram illustrating the overlap of frailty, low muscle strength, and low muscle mass. The dashed line frames the group of patients with co-occurrence of sarcopenia and frailty in older mobility-limited patients.

The mean HGS in the total population was 23.4 ± 9.2 kg (males: 32.4 ± 8.4 vs females: 18.6 ± 5.2 kg, p<0.001), with low HGS present in 28% of the overall group of patients (males: 23.8% vs females: 30.2%, p=0.192). At the same time, STS performance was 9.7 ± 5.2 repetitions (males: 9.8 ± 6.1 vs females: 9.6 ± 4.7, p=0.545) and 36.8% had low STS performance (males: 36.9% vs females: 36.7%, p=0.971) (Table 2).

Low muscle strength (low HGS and/or low STS) was present in 50.9% (males: 48.5% vs females: 52.2%, p=0.516) of the total cohort, while the co-prevalence of low HGS and low STS performance was 13.9% (males: 12.3% vs females: 14.7%, p=0.525).

Mean SMI for the entire cohort was 6.6 ± 1.1 kg/m^2^ (males: 7.6 ± 1.0 vs females: 6.1 ± 0.8 kg/m^2^, p<0.001) with low SMI present in 25.1% (males: 27.7% vs females: 23.7%, p=0.393).

The mean habitual GS was 0.87 ± 0.26 m/sec (males: 0.89 ± 0.25 vs females: 0.86 ± 0.26, p=0.398), with low habitual GS present in 40.3% (males: 33.9% vs females: 43.8%, p=0.065). The mean maximal GS was 1.16 ± 0.37 m/sec (males: 1.23 ± 0.38 vs females: 1.13 ± 0.35, p=0.028) with low maximal GS present in 16.3% (males: 13.4% vs females: 18%, p=0.270).

## 4. Discussion

In the present study, we examined sarcopenia, frailty, and their individual components in a cohort of mobility-limited older adults using both screening instruments and consensus-based diagnostic criteria. By contrasting risk-based screening with confirmed diagnoses, the study aimed to clarify how different assessment approaches capture distinct aspects of muscle-related vulnerability.

Within this framework, low muscle mass was observed in 25.1% of the patients. In contrast, nearly half of the cohort (48.0%) was classified as being at risk of sarcopenia by SARC-F, whereas only 14.1% met the EWGSOP2 diagnostic criteria. Frailty was identified in 23.7% of patients, however the co-prevalence of frailty and sarcopenia was limited, with only 7.2% of patients presenting with both conditions simultaneously. This distinction between frailty and sarcopenia underscores the complexity of managing these conditions in mobility-limited older individuals.

While approximately 25% of the patients could be characterized as frail and with low muscle mass, more than 50% demonstrated low muscle strength. In addition, the low co-prevalence (13.9%) of low HGS and STS performance highlights the complex and multifaceted nature of skeletal muscle impairment in this distinct patient population.

The discrepancy between the 48.0% of patients identified as potentially having sarcopenia by the SARC-F questionnaire and the 14.1% who fulfilled the EWGSOP2 criteria for sarcopenia highlights important considerations in screening and diagnosis. The SARC-F questionnaire is a simple and widely used tool designed to identify individuals at risk for sarcopenia based on self-reported functional limitations. However, its high sensitivity may lead to overestimation of sarcopenia prevalence, as seen in this study, where nearly half of the patients were identified as at risk. These results underscore the importance of using a multi-step approach in sarcopenia diagnosis, where initial screening tools like SARC-F should be followed by confirmatory assessments to ensure accurate diagnosis and appropriate management of sarcopenia in older adults.

In the present study, the prevalence of sarcopenia and severe sarcopenia was 14.1% and 8.3%, respectively, according to the EWGSOP2 diagnosis criteria (Cruz-Jentoft et al., 2019). This finding is consistent with a Korean study, which reported the prevalence of sarcopenia and severe sarcopenia to be 18.4% and 3.9%, using the EWGSOP2 criteria, in 2099 ambulatory community-dwelling older adults (Kim & Won, 2019). Notably, the prevalence of sarcopenia not only depends on the population, but also the criteria used. Hence, in a recent meta-analysis (Petermann-Rocha et al., 2022), the prevalence of sarcopenia varied from 10% (95% CI: 2.0–17.0%, EWGSOP2 (Cruz-Jentoft et al., 2019)) to 22% (95% CI: 22.0–25.0%, EWGSOP (Cruz-Jentoft et al., 2010)) depending on the EWGSOP criteria, reflecting the importance of the used, classification systems, cut-off points, and diagnostic tools to measure muscle mass (Mayhew et al., 2019; Petermann-Rocha et al., 2022).

The observed frailty prevalence of 23.7% is consistent with prior reports. A scoping review showed that the prevalence of frailty evaluated by the CFS ranges from 25-48% among geriatric outpatients (Church et al., 2020). The higher prevalence of frailty compared to sarcopenia in the present cohort could suggests that frailty reflects a broader range of physiological impairments beyond muscle mass and strength, including cognitive decline, nutritional deficiencies, and comorbid conditions.

The co-prevalence of frailty and sarcopenia of 7.2% in the present population indicates an overlap, although minor in magnitude, between these two conditions. This suggests that there is a subset of older adults who experience compounded vulnerabilities, with both muscle impairments and systemic physiological decline, in turn contributing to functional limitations such as increased fall risk and reduced mobility. The concurrence of these conditions exacerbates the risk of negative health outcomes, as individuals with both sarcopenia and frailty are likely to have even greater impairments in mobility, balance, and overall physical function. These findings have important clinical implications for the management of older mobility-limited patients, by stressing the need for assessing both sarcopenia and frailty in this population, while also pointing to the necessity for multifaceted intervention strategies that address both muscle strength and broader physiological resilience.

For older people, muscle strength is essential for performing daily activities such as walking, climbing stairs, and lifting objects as well as supporting vertical posture and postural balance (Chodzko-Zajko et al., 2009). According to EWGSOP2, muscle strength is also considered the primary indicator of sarcopenia (Cruz-Jentoft et al., 2019). In the present study, approximately 50% of the patients had low muscle strength, as indicated by either low HGS and/or low STS performance. Specifically, 28% of the patients had low HGS, while ∼37% had low STS performance. The reported mean HGS values (∼32 kg in males and ∼19 kg in females) are in line with previous findings in similar populations with values ranging from 30-33 kg in males and 18-21 kg in females (Barnes et al., 2018; Christensen et al., 2018; Kim & Won, 2019).

Notably, however, even though approximately 50% of the patients had low muscle strength, only 13.9% demonstrated both low HGS and low STS performance, indicating only a small degree of overlap between age-related impairments in upper and lower body muscle function. Comparable findings have been reported previously among ambulatory community-dwelling older adults (Kim & Won, 2019), with 23.5% exhibiting low muscle strength by either low HGS (13.7%) or low STS performance (13.6%), while only 3.5% demonstrated both low HGS and STS performance (Kim & Won, 2019). These findings suggest that it becomes of importance to assess both HGS and STS performance in older individuals to identify conditions of diminished muscle strength and not overlook the risk of sarcopenia.

Among the different assessments, STS performance was the only with no difference between sexes. The STS test is one of the few strength/power-focused assessments where both males and females typically exhibit similar performance levels. This phenomenon is likely attributed to the fact that females, on average, tend to have shorter stature than males, which make it easier for them to rise from a seated position. Conversely, males are typically taller and in general characterized by greater muscle strength compared to females. Also, the STS test requires the handling and transfer of one’s body weight, inherently incorporating a body weight normalization factor. As a result, despite the inherent differences in strength between sexes, the STS test provides a relatively equitable measure of lower body muscle strength/power across both sexes (Alcazar et al., 2021). This characteristic makes the STS test a valuable tool for assessing mechanical muscle function in various populations, regardless of sex-differences.

In the present study, 25.1% of patients were found to have low muscle mass, yet only 14.1% met the criteria for sarcopenia. This discrepancy suggests that there may be individuals with low muscle mass who do not meet the diagnostic criteria for sarcopenia, potentially leading to underdiagnosis of sarcopenia/muscle-related abnormalities. This highlights the importance of assessing muscle mass in all patients, regardless of whether muscle strength is normal or reduced, in order to intervene early and prevent further muscle decline, thereby reducing the risk of an increase in the proportion of older adults developing sarcopenia.

Nonetheless, in this study, we found a mean SMI of 6.6 kg/m^2^, which is in line with other studies involving similar patient groups (Christensen et al., 2018; Kim & Won, 2019). Importantly, comparing absolute SMI values across studies is challenging due to the use of different measurement methods, such as BIA and DXA, which may yield varying results, making direct comparisons difficult. Additionally, the cut-off values for low SMI varies depending on the specific diagnostic criteria used, which further complicate comparisons and affecting the reported prevalence rates (Van Ancum et al., 2020). However, the observed values were slightly lower compared to those reported in studies of healthy community-dwelling older people (Alcazar et al., 2018; Bahat et al., 2021). Alcazar et al (Alcazar et al., 2018) reported a mean SMI of 7.2 kg/m^2^, while Bahat et al (Bahat et al., 2021) reported an SMI of 10.2 kg/m^2^, although the sex distribution was largely similar across all the studies (including the present cohort) (Alcazar et al., 2018; Bahat et al., 2021; Christensen et al., 2018; Kim & Won, 2019). Hence, the variability in skeletal muscle mass between mobility-limited geriatric outpatients and community-dwelling older adults may be attributed to reduced physical activity, underlying health conditions, insufficient nutrition, medication side effects, and/or elevated levels of low-grade inflammation and oxidative stress (Tieland et al., 2018).

These observations indicate that low muscle strength and low muscle mass do not necessarily concur within the same individual underscoring the complexity of muscle health in old age and stresses the multifactorial nature of frailty and sarcopenia. Potential reasons exist for older adults to exhibit low muscle strength without a corresponding decrease in muscle mass, and vice versa. Muscle strength and power are influenced by both muscular and neural factors, such as neuromuscular activation and muscle quality, while muscle mass primarily reflects the quantity of tissue. Thus, individuals may have sufficient muscle mass but reduced strength due to factors like poor neuromuscular activation or muscle quality (Aagaard et al., 2010; Goodpaster et al., 2006). Conversely, older adults with low muscle mass may not necessarily exhibit reduced muscle strength if the muscles are efficiently activated and functionally adapted. In support of the latter notion, lifelong physical activity is known to have a positive effect on muscle strength even in the presence of low muscle mass (Ramsey et al., 2021). Additionally, factors such as hormonal status, nutrition, and chronic diseases further contribute to discrepancies between muscle mass and strength (Larsson et al., 2019). Therefore, understanding the complex interplay between muscle mass, strength, and various influencing factors is essential for accurately diagnosing and managing conditions such as sarcopenia and optimizing muscle health across the lifespan.

### 4.1. Limitations and strengths

The present study has both certain strengths and limitations.

One limitation is the cross-sectional study design, so any day-to-day variance in physical performance was not accounted for within this design. Another limitation is the hydration status of the patients could impact the result of the bioelectrical impedance analysis, although only limb muscle mass was assessed by SMI, thereby reducing a major part of the variance due to hydration status.

However, the present study also has several strengths. One of the strengths is the relatively large sample size. Given the comprehensive nature of the examinations and assessments conducted, the number of patients included is relatively substantial. Another strength is that the present study is the first, at least to our best knowledge, to examine the prevalence of sarcopenia in a high-risk population of mobility-limited geriatric patients using the diagnostic criteria proposed by EWGSOP2. Finally, we measured both HGS and STS performance to identify patients with low upper and lower body muscle strength/power to ensure individuals with deficits in one parameter were not overlooked, thereby optimizing the detection of muscle weakness.

## 5. Conclusion

In conclusion, sarcopenia was present in 14.1% of mobility-limited older adults according to the EWGSOP2 criteria, whereas frailty was identified in 23.7% of the cohort. Despite this, the co-prevalence of frailty and sarcopenia was limited, with only 7.2% of patients exhibiting both conditions. Moreover, approximately 25% of patients demonstrated low muscle mass, whereas the co-prevalence of low HGS and low STS performance was uncommon.

These findings indicate that low muscle mass and impaired muscle function do not necessarily coexist, suggesting that reliance on a single domain may lead to under-recognition of sarcopenia or other clinically relevant muscle impairments. Accordingly, the results underscore the need for a comprehensive assessment strategy incorporating both upper- and lower-body muscle function alongside muscle mass. Given the higher prevalence of frailty relative to sarcopenia, systematic evaluation of both conditions is essential in mobility-limited older adults to enable targeted, multidimensional intervention strategies.

## Data Availability

The datasets generated and/or analyzed during the current study are not publicly available due data privacy concerns and ethical restrictions but are available from the corresponding author (PH) upon reasonable request.

## List of abbreviations

ADL: Activities of daily living,
ALM: Appendicular Lean Mass,
BMI: Body Mass Index,
CFS: Clinical Frailty Scale,
BIA: Direct Segmental 8-electrode Multi-frequency Bioelectrical Impedance Analyses,
DXA: Dual-energy X-ray Absorptiometry,
EWGSOP: European Working Group on Sarcopenia in Older People,
GDS-15: Geriatric Depression Scale,
GS: Gait Speed,
HGS: Hand grip strength,
Kg: Kilograms,
M: Meter,
SD: Standard Deviation,
SMI: Skeletal Muscle Index,
SNAQ: The Short Nutritional Assessment,
STS: 30 second Sit-to-Stand.

## Declarations

### Ethics approval and consent to participate

All enrolled patients provided written consent after being informed about the study. The study was approved by the local Ethics Committee of Copenhagen and Frederiksberg (H-20057620), the Danish Data Protection Agency (P-2021-619), and registered at Clinicaltrials.gov (NCT05795556).

### Consent for publication

Not applicable. This manuscript does not contain any individual person’s data in any form. All patients were anonymized using ID numbers during the analysis.

### Funding

The work was supported by funding from the Research Council at Herlev and Gentofte Hospital, the Tømmerhandler Johannes Fog Foundation, and the Aase and Ejnar Danielsens Foundation.

### CRediT authorship contribution statement

**PH:** Conceptualization, Methodology, Investigation, Data curation, Formal analysis, Project administration, Visualization, Writing – original draft. **HN:** Conceptualization, Supervision, Formal analysis, Data curation, Writing – review and editing. **RSK:** Conceptualization, Methodology, Writing – review and editing. **MS:** Writing – review and editing. **JR:** Writing – review and editing. **PAA:** Writing – review and editing. **FD:** Resources, Writing – review and editing. **CS:** Conceptualization, Methodology, Supervision, Writing – review and editing.

### Competing interests

PH, HN, RSK, MS, JR, PA, FD, and CS declare that they have no conflict of interest.

## Acknowledgements

The authors extend their gratitude to the physiotherapist, doctors, nurses, and secretaries at the outpatient clinic at Herlev and Gentofte Hospital for their cooperation and patience during patient recruitment and assessment. Additionally, special thanks are extended to the physiotherapists for their assistance in patient assessment.

## Notes

### Competing Interest Statement

The authors have declared no competing interest.

### Author Declarations

The Regional Committee on Health Research Ethics of the Capital Region of Denmark granted ethical approval for this study (H-20057620).

